# Development of a SARS-CoV-2 total antibody assay and the dynamics of antibody response over time in hospitalized and non-hospitalized patients with COVID-19

**DOI:** 10.1101/2020.06.17.20133793

**Authors:** Erik H. Vogelzang, Floris C. Loeff, Ninotska IL Derksen, Simone Kruithof, Pleuni Ooijevaar-de Heer, Gerard van Mierlo, Federica Linty, Juk Yee Mok, Wim van Esch, Sanne de Bruin, Alexander P.J. Vlaar, AmsterdamUMC COVID-19 biobank study group, Bart Seppen, Maureen Leeuw, Anne J.G. van Oudheusden, Anton G.M. Buiting, Kin Ki Jim, Hans Vrielink, Francis Swaneveld, Gestur Vidarsson, C. Ellen van der Schoot, Peter Wever, Wentao Li, Frank van Kuppeveld, Jean-Luc Murk, Berend Jan Bosch, Gerrit-Jan Wolbink, Theo Rispens

**Author notes:** contributed equally (Erik H. Vogelzang, Floris C. Loeff). see appendix for author list of the study group.

## Abstract

SARS-CoV-2 infections often cause only mild disease that may evoke relatively low antibody titers compared to patients admitted to hospitals. Generally, total antibody bridging assays combine good sensitivity with high selectivity. Therefore, we developed sensitive total antibody bridging assays for detection of SARS-CoV-2 antibodies to the receptor-binding domain (RBD) and nucleocapsid protein (NP), in addition to conventional isotype-specific assays. Antibody kinetics was assessed in PCR-confirmed hospitalized COVID-19 patients (n=41) and three populations of patients with COVID-19 symptoms not requiring hospital admission: PCR-confirmed convalescent plasmapheresis donors (n=182), PCR-confirmed hospital care workers (n=47), and a group of longitudinally sampled symptomatic individuals highly suspect of COVID-19 (n=14). In non-hospitalized patients, the antibody response to RBD is weaker but follows similar kinetics as has been observed in hospitalized patients. Across populations, the RBD bridging assay identified most patients correctly as seropositive. In 11/14 of the COVID-19-suspect cases, seroconversion in the RBD bridging assay could be demonstrated before day 12; NP antibodies emerged less consistently. Furthermore, we demonstrated the feasibility of finger prick sampling for antibody detection against SARS-CoV-2 using these assays. In conclusion, the developed bridging assays reliably detect SARS-CoV-2 antibodies in hospitalized and non-hospitalized patients, and are therefore well-suited to conduct seroprevalence studies.

## Introduction

In December 2019, the first cases of atypical pneumonia were reported in China.[1] The causative agent was a β-coronavirus; now known as severe acute respiratory syndrome coronavirus 2 (SARS-CoV-2). SARS-CoV-2 is related to the human severe acute respiratory syndrome coronavirus 1 (SARS-CoV-1) and BatCoV RaTG13.[2] In the following months, an increase in reverse transcription-polymerase chain reaction (RT-PCR) confirmed cases of coronavirus infectious disease 2019 (COVID-19) were reported in various countries resulting in the World Health Organization (WHO) declaring a pandemic. RT-PCR-based diagnostics requires active virus shedding. As such, many COVID-19 patients that were not hospitalized remain undiagnosed, and the true extent of SARS-CoV-2 infections is unquestionably higher than initially reported.[3,4] This is confirmed by several seroprevalence studies that have since been conducted.[5–7] However, insight into the dynamics and variability of the antibody response in non-hospitalized patient populations, with only mild symptoms remains to be elucidated. Furthermore, the suitability of various assays to assess these – presumably partly rather weak – responses, is a point of concern.

The coronavirus comprises four common structural proteins; the spike (S), envelope (E), membrane (M) and nucleocapsid (N) proteins.[8,9] Previous studies on SARS-CoV-1 identified the S protein as a major antigen and antibodies against the S protein were demonstrated to have neutralizing capacity.[10–14] The S protein and more specifically the receptor-binding domain (RBD) in subunit S1 in SARS-CoV-2 is reported to be an immunodominant epitope [40], and has been demonstrated to have negligible cross-reactivity with human coronaviruses HKU1, 229E, NL63 and OC43 or Middle East Respiratory Syndrome coronavirus (MERS-CoV) and only to SARS-CoV-1.[15] The N protein (NP) in SARS-CoV-1 is also immunogenic, eliciting an antibody response.[16,17]. Studies on SARS-CoV-1 and SARS-CoV-2 reported that antibodies against the N protein appeared earlier than those directed at the S protein.[18–22] Furthermore, in SARS-CoV-2, immune-dominance of the N-antigen in COVID-19 patients was described.[23] Implementing validated serological assays will provide much-needed information on transmission, the ascertainment of the true infection rate, corresponding mortality and morbidity and the immune response. Also, it will allow a better understanding of the dynamics of SARS-CoV-2 in the general population.

Here, we describe the development of a serological bridging assay for the detection of antibodies against the RBD and NP of SARS-CoV-2. Bridging assays, or double-antigen sandwich assay, have the advantage of being able to detect antibodies regardless of isotype, which potentially makes these assays maximally sensitive for seroprevalence studies. Here, we used these assays, together with isotype-specific assays, to assess the antibody response in several hospitalized and non-hospitalized COVID-19 patient populations. In particular, we describe the early antibody response to SARS-CoV-2 in non-hospitalized individuals highly suspect of COVID-19, during the symptomatic stages of disease.

## Materials and Methods

### Samples and patients

Serum or plasma samples obtained from RT-PCR positive SARS-CoV-2 hospitalized and non-hospitalized recovered adult patients (n=182) who underwent plasmapheresis at Sanquin, the national blood bank, Amsterdam, the Netherlands were included.

Also, serum samples from hospitalized SARS-CoV-2 RT-PCR confirmed patients (n=41) at the Jeroen Bosch Hospital, Den Bosch, the Netherlands and Amsterdam University Medical Centre, Location AMC, Amsterdam, the Netherlands were included.

From the Elisabeth-TweeSteden Hospital, Tilburg, the Netherlands serum samples of SARS-CoV-2 RT-PCR confirmed healthcare workers (HCWs) were obtained (n=47) at various time points. None of the HCWs were admitted to the hospital for COVID-19.

From the Amsterdam Rheumatology and immunology Centre, location Reade, The Netherlands, serum samples were obtained at regular intervals in a group of otherwise healthy volunteers which based on recent travel to an endemic area abroad were highly suspect for COVID-19 (n=7) and household contacts (n=2) of this group that did not go abroad. Moreover, serum samples were collected from a SARS-CoV-2 RT-PCR positive volunteer and close contacts, all with no comorbidities. (n=5). All 14 volunteers had symptoms not requiring hospital admission. Samples were collected at regular intervals after the onset of potential COVID-19 disease symptoms. The average age of participants was 31 years (SD 4.2), 7 (50%) were female and none reported relevant comorbidities.

Negative control serum samples were included from healthy blood donors collected by Sanquin (n=307). The negative control samples had been collected from the Dutch population before mid-January 2020, thus prior to the first reported SARS-CoV-2 case in the Netherlands (27 February 2020).

This study has been conducted in accordance with the ethical principles set out in the declaration of Helsinki and all participants provided written informed consent, if applicable. Approval was obtained from the Medical Ethics Committees from the Academic Medical Center, VU University Medical Center and Elisabeth-TweeSteden Hospital.

### Development of SARS-CoV-2 RBD and NP based bridging assays

The RBD-mFc and RBD-ST proteins were produced as described by Okba et al.[15] The NP was produced in HEK cells with HAVT20 leader peptide, 10xhis-tag and a BirA-tag as described by Dekkers et al.[24] MaxiSORP microtiter plates (ThermoFisherScientific, US) were coated with 100 µL per well 0.125 µg/mL RBD-mFc or NP in PBS overnight at 4 ° C. Plates were washed five times with PBS supplemented with 0.02% polysorbate-20 (PBS-T). Samples were diluted 10-fold in PBS-T supplemented with 0.3% gelatin (PTG) and incubated on the plate (100 µL) for 1h at room temperature (RT) while shaking. After washing five times with PBS-T, 100 µL per well biotinylated RBD-ST (EZ-Link Sulfo-NHS-LC-Biotin, Thermo Fisher) or biotinylated NP (via the BirA tag as described in [24]) was added at 0.5 µg/mL or 0.015 µg/mL, respectively, in PTG and incubated for 1h at RT, followed by incubation for 30 minutes with streptavidin-poly-HRP (Sanquin). Plates were washed five times with PBS-T, and 100 μl of TMB substrate (100 μg/mL) and 0.003% (v/v) hydrogen peroxide (Merck, Germany) in 0.11 M sodium acetate buffer (pH 5.5) was added to each well. A total of 100 μl of 0.2 M H_2_SO_4_ (Merck, Germany) was added to stop the reaction. Absorbance was measured at 450 nm and 540 nm. The difference was used to evaluate antibody binding. OD values were normalized to readings of a reference serum pool that was included on each plate and reported as nOD.

### SARS-CoV-2 RBD and NP isotype-specific assays

MaxiSORP microtiter plates were coated with 1.0 µg/mL RBD-ST or NP, or 4.0 µg/mL monoclonal mouse antihuman IgM (MH15, Sanquin, the Netherlands) in PBS overnight at 4 ° C. Following washing with PBS-T, samples were diluted 100-fold/200-fold in PTG and incubated for 1h at RT. After washing, 0.5 µg/mL HRP-conjugated monoclonal mouse antihuman IgG (MH16, Sanquin) or antihuman IgA (MH14, Sanquin) was added, diluted in PTG and incubated for 1h. Following enzymatic conversion of TMB substrate, absorbance was measured at 450 nm and 540 nm and the difference used to evaluate antibody binding as described above. For IgM, the assay was finished using biotinylated RBD-ST as described above for the bridging assay.

### Data processing and analysis

Statistical analysis were carried out using Graphpad Prism 7.

## Results

### Total antibody assays

Total antibody bridging assays depend on the multivalency of antibodies of any isotype to generate a bridge between antigen for capture as well as detection. Therefore, antigen coating density is generally a critical parameter. We set up bridging assays for RBD and NP antigens and optimized antigen coating density to obtain optimal differentiation between specific and non-specific signals using a small set of sera from COVID-19 patients and pre-outbreak healthy donors (HD), see Figure S1. A good reproducibility between runs was observed (Figure S1). After an initial evaluation of 80 pre-outbreak samples, we arrived at a cut-off value of 0.1 nOD, anticipated to provide ca. 99% specificity. Subsequent samples were measured, retested if initially positive (>0.1 nOD), and only considered positive if the repeated measurement was also >0.1 nOD. Analogously, a cut-off value of 0.14 nOD was set for the NP bridging assay.

### Seroprevalence in different study populations

We assessed seroprevalence with the RBD bridging assay (RBD-Ab) and NP bridging assay (NP-Ab) in several different study populations of PCR-confirmed COVID-19 patients as well as pre-outbreak HD. Only two (0.7%) positive samples were detected in pre-outbreak HD (n=307) using the RBD-Ab, whereas seven (2.5%) positive samples were detected using the NP-Ab (Figure 1). In 40 out of 41 (97.5%) and 39 out of 41 (95.1%) hospitalized patients with active, RT-PCR confirmed COVID-19 we detected antibodies against the RBD or NP, respectively. In PCR-confirmed non-hospitalized convalescent HCW’s (n=47) antibodies against RBD or NP were detected in 45 (95.7%) and 39 (83.0%) samples, respectively. Antibodies against RBD were detected in 180 (98.9%) and for the NP in 175 (96.2%) of PCR-confirmed convalescent plasmaphereses donors (n=182), which consists both of patients with and without prior hospitalization due to COVID-19. In all but one case, samples negative in the RBD-Ab assay were also negative in the NP-Ab assay (Figure 4). The above-described seroprevalences demonstrated a good overall sensitivity and specificity of the RBD-Ab (98.1% and 99.3%), and somewhat less for the NP-Ab (93.7% and 97.5%).

**Figure 1.**
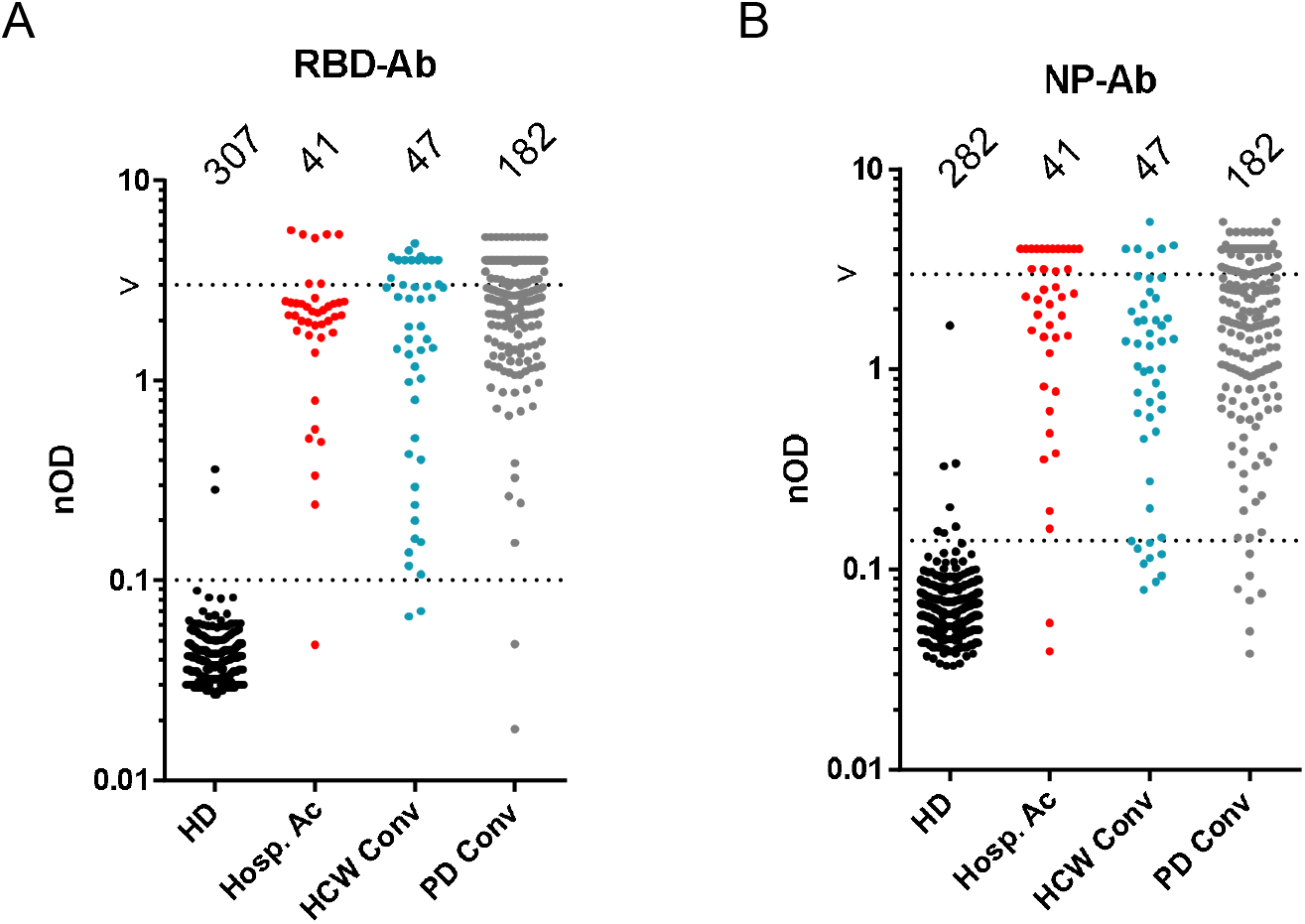
Total antibody titers measured in different patient populations. Total antibody titers against the RBD (A) and NP (B) measured in different patient populations: pre-outbreak healthy donors (HD), hospitalized active-disease patients (Hosp. Ac), non-hospitalized, convalescent healthcare workers (HCW Conv) and convalescent plasmapheresis donors (PD Conv). Samples below the lower dotted line are considered negative, whereas the upper dotted line indicates upper boundary of dynamic range.

**Figure 4.**
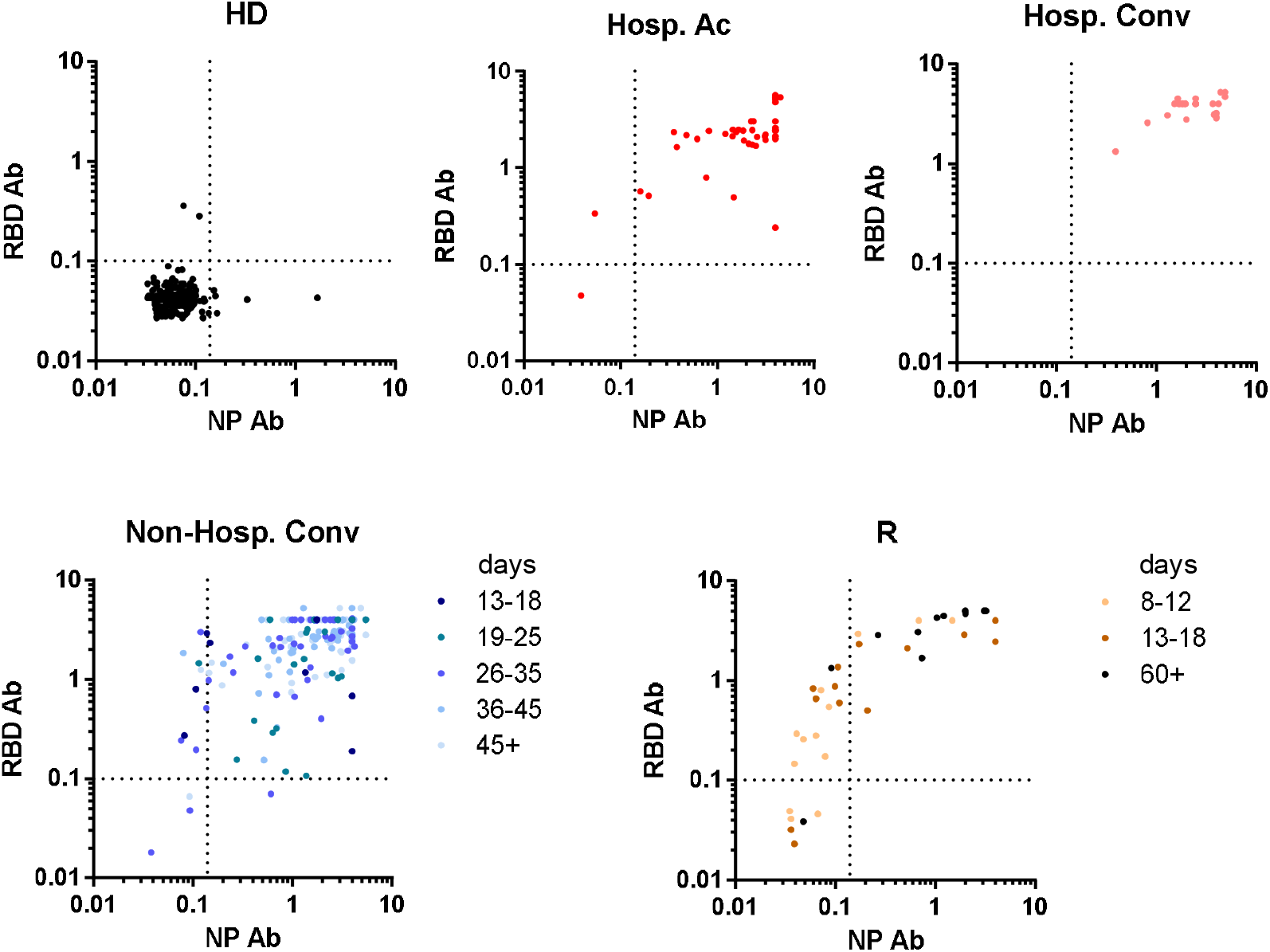
Anti-NP and RBD immune responses compared. Antibodies detected with the NP-Ab and the RBD-Ab assay plotted on the x-y axes for different patients groups: pre-outbreak healthy donors (HD), hospitalized patients with active disease(Hosp. Ac) or convalescent cases (Hosp. Conv), non-hospitalized convalescent individuals (Non-Hosp. Conv, and the Reade patient cohort (R); stratified according to days post symptom onset as indicated. Dotted lines indicate the cut-off for both assays.

### Antibody response at different time points

To study the effect of time on the seroprevalence and antibody levels determined by using the RBD-Ab and the NP-Ab, we stratified the samples of the non-hospitalized cases (plasmapheresis donors and HCWs) for different days since symptom onset (information available for n=167; Figure 2). As comparison, in addition to the hospitalized patients with active disease (approx. 8 – 23 day post symptom onset), convalescent cases that were previously admitted to the hospital are also included (n = 27; approx. 20 – 66 post symptom onset). There is an increase in nOD over time for the RBD-Ab in hospitalized cases and a similar, non-significant trend in non-hospitalized cases. However, there is no apparent effect of time after onset of symptoms on the detection of antibodies in the RBD-Ab or NP-Ab assay.

**Figure 2.**
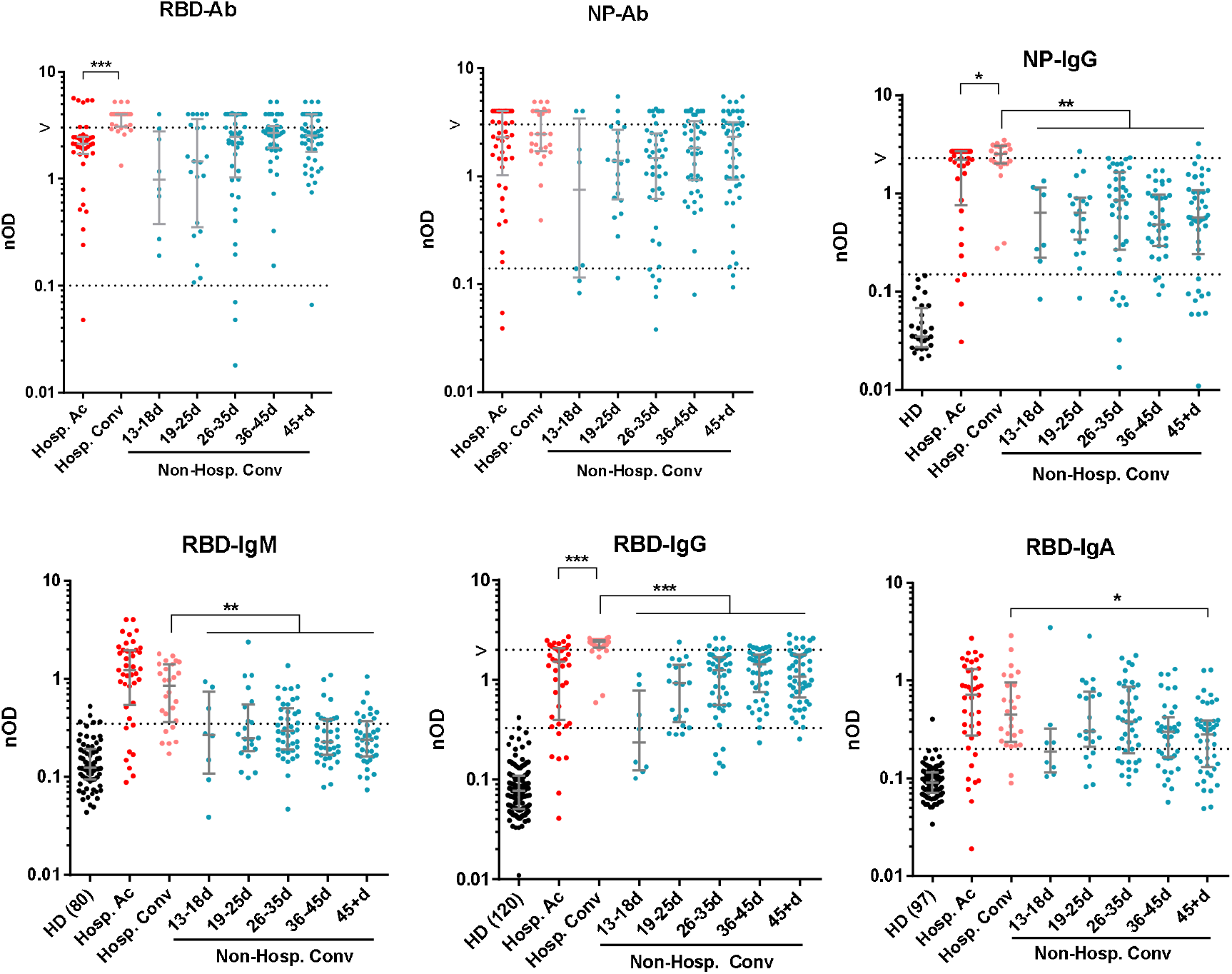
Antibody titers and isotypes measured in different patient populations and at various time points. Total and isotype-specific antibody titers against the RBD and NP were measured in different patient populations: hospitalized patients with active disease (Hosp. Ac; 8 – 23 days post symptom onset; n = 41) or convalescent cases (Hosp. Conv; 20 – 66 days post symptom onset; n = 27), and non-hospitalized convalescent individuals (Non-Hosp. Conv; n = 167). For the latter group, on the x axes the number of days post symptom onset are indicated. For the isotype-specific assays, signals measured in pre-outbreak healthy donors (HD) are also indicated (for total Ab, see Figure 1). Dotted lines indicate approximate 95% (IgM) and 98% (IgG, IgA) percentile of the negative controls. * p < 0.05; ** p < 0.01; *** < 0.001. Tests performed: Hosp. Ac vs Hosp. Conv, Mann-Whitney; Hosp. Conv vs Non-Hosp. Conv, Kruskal-Wallis test/Dunn’s multiple comparisons test.

To obtain insight into the dynamics of the different isotypes, we also measured antibodies using assays for the detection of IgM, IgG, and IgA (Figure 2). For the antibody response to the RBD, there is a (non-significant) trend towards declining IgM and IgA levels as time progresses in both hospitalized and non-hospitalized patients. Hospitalized patients have significantly higher IgG levels later in time. In non-hospitalized patients, the IgG response rises slowly over time and gains prominence relative to other isotypes at later time points (Figure S2), in line with expectations. At least up to 60 days after symptom onset substantial amounts of IgG to the RBD can be detected in this population. In comparison to hospitalized convalescent patients, the IgG response in non-hospitalized patients is significantly weaker, as shown in Figure 2. A similarly weaker IgG response is also observed against the NP.

### Antibody kinetics in non-hospitalized patients at early time points

To obtain a better insight into the antibody response against SARS-CoV-2 and thus the seroprevalence in a in specific population, we tested sequentially collected sera from a group of non-hospitalized and otherwise healthy individuals who were highly suspected of having contracted COVID-19 (Reade patient cohort; Figure 3). Day 3 post symptom onset samples were available for two individuals and were negative in both the bridging assays as well as in the IgM, IgG, and IgA assays. Within 12 days post symptom onset, antibodies against the RBD were detected in 11 out of 14 (78.6%) patients as compared to 8 out of 14 (53.1%) against NP antibodies, as shown in Figure 3, using the bridging assays. At day 60, one of the previously negative samples became positive in both bridging assays.

**Figure 3.**
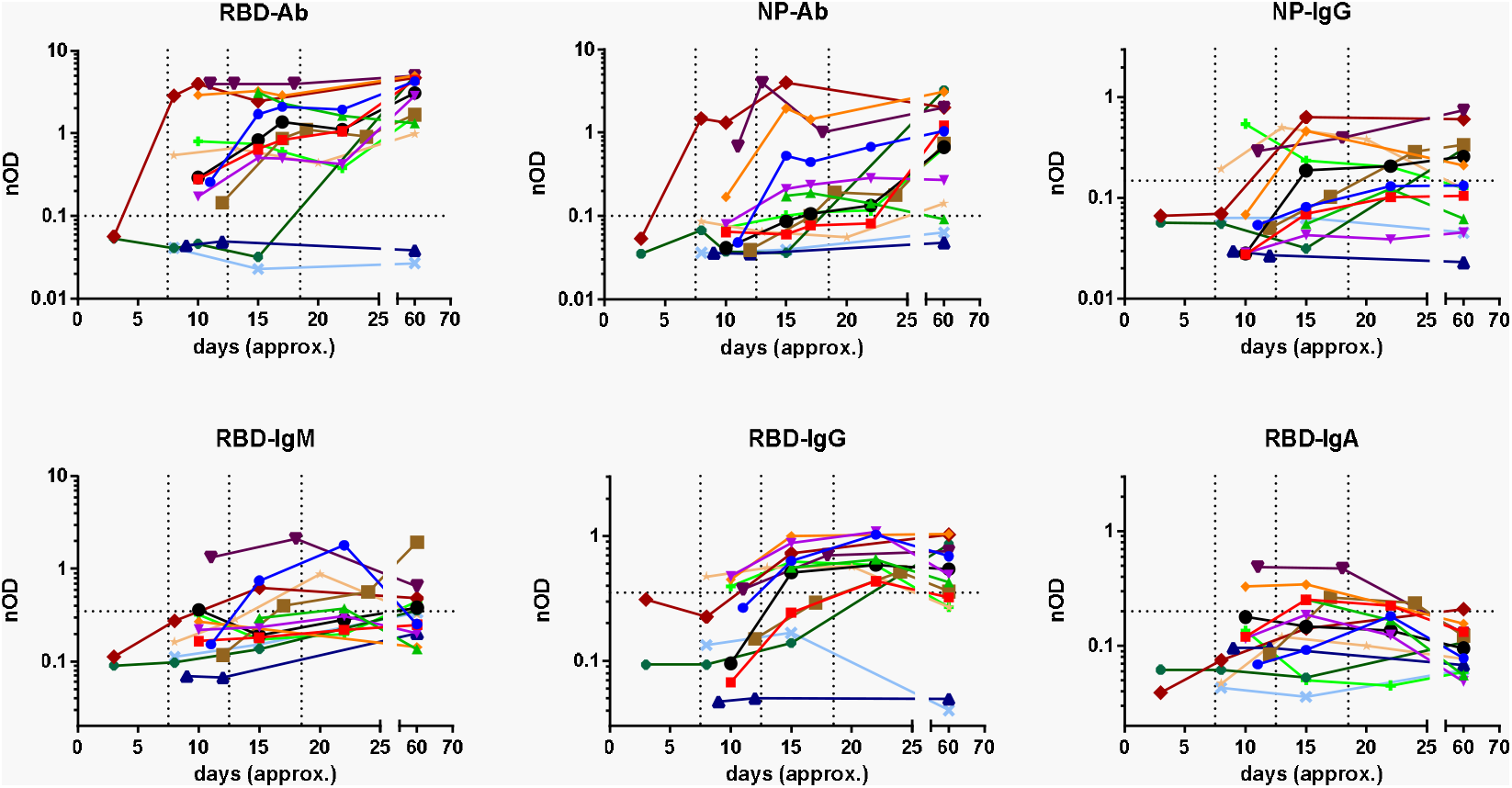
Kinetics of antibody response against SARS-CoV-2. In 14 non-hospitalized patients with mild symptoms and a high likelihood (1 SARS-CoV-2 RT-PCR confirmed) of SARS-CoV-2 infection we tested the antibody response against the RBD-Ab, NP-Ab, NP-IgG, RDB-IgM, RBD-IgG and RBD-IgA. The x-axes show the number of days post symptom onset. The horizontal dotted line is the cut-off; samples below this line are considered negative. The vertical dotted lines represent from left to right 8 days, 13 days and 19 days post symptom onset.

Figure 3 shows the antibody kinetics for IgM, IgG and IgA. Most patients (n=9) seroconverted for IgG within 15 days post symptom onset, although three patients seroconverted later. The slow appearance of IgG antibodies to RBD is clearly seen in this population. Interestingly, at two months, IgG anti-RBD appears to be declining again, suggesting a lack of sustained overall anti-RBD response in this group of patients. The antibody response to NP is not detectable in most patients around day 10 both in the NP-Ab and NP-IgG assays, and remains weak in many patients at later time points.

### Comparison of assay formats

The RBD-Ab assay identifies positive patients most reliably, with few false-positive cases. Sensitivity and/or specificity could theoretically be enhanced by combining two or more assays. When comparing the RBD-Ab with the NP-Ab assay, we observe that for HD, none of the samples were positive in both assays as shown in Figure 4. Requiring both assays to be positive would therefore result in enhanced specificity, albeit with reduced sensitivity. Also, for the RBD-IgG assay it appears that negative control samples either have elevated signals in either this assay or the RBD-Ab, but not both (Figure 5). There is a moderately strong correlation between RBD-Ab and RBD-IgG (Spearman r = 0.69, p < 0.0001) across the populations; whereas the correlation between RBD-Ab and NP-Ab is moderate (r = 0.58, p<0.0001).

**Figure 5.**
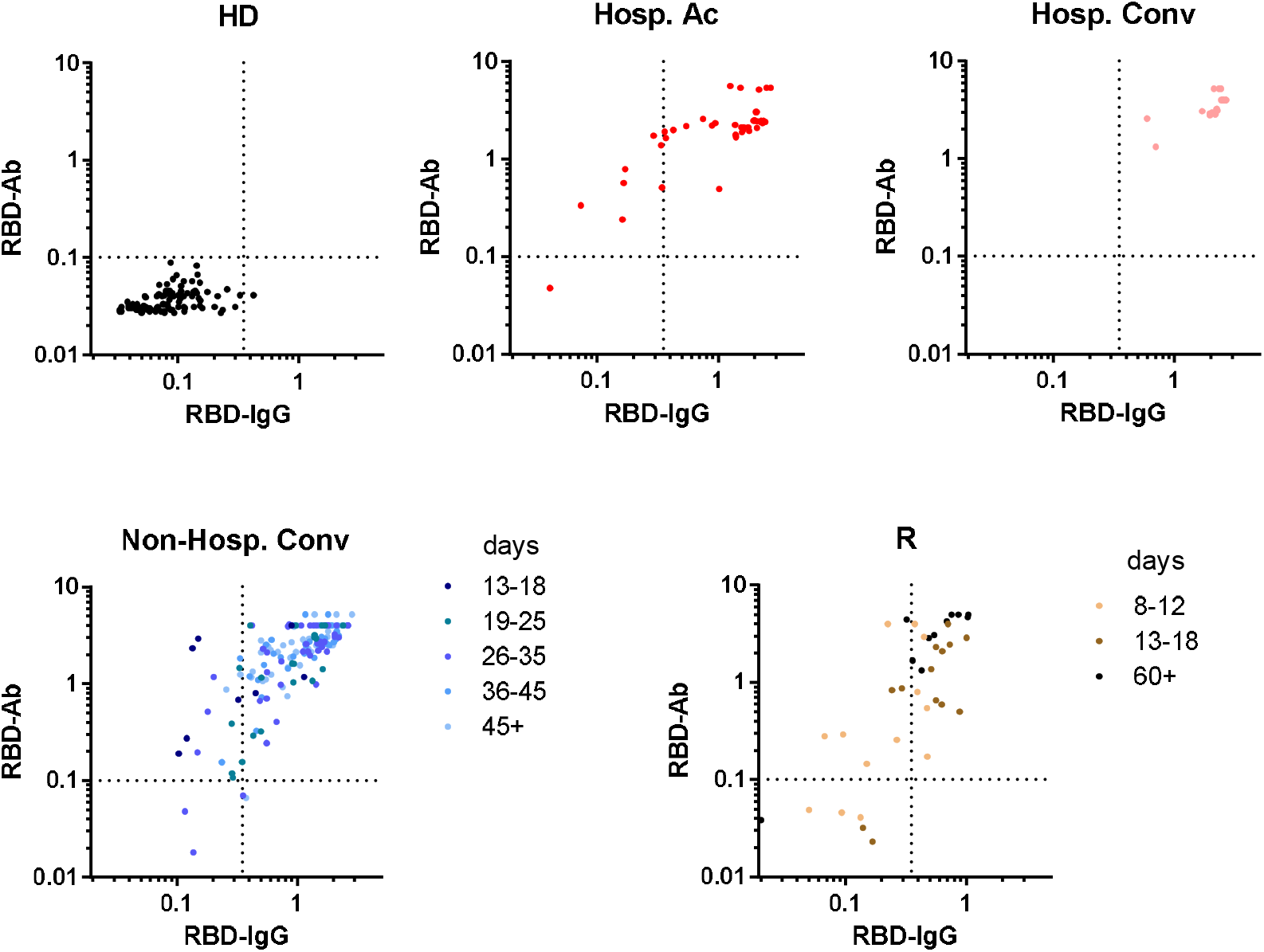
Comparison between the RBD-Ab and RBD-IgG assay. Comparison of the detection of antibodies with the RBD-Ab and RBD-IgG assay plotted on the x-y axes for different patient groups; stratified according to days post symptom onset as indicated. Dotted lines indicate the cut-off for both assays.

### Finger prick sampling

A relatively low volume of serum is needed for the developed assay to assess the antibody response, therefore making it well suited for finger prick sampling. A small number of samples (n = 11) was obtained by finger prick sampling, rather than obtained by venipuncture. Antibodies were detected by the RBD-Ab assay and compared to a matching venipuncture draw (Figure 6). The results suggest that a finger prick is an equivalent method for drawing blood when using this assay to detect antibodies against the RBD.

**Figure 6.**
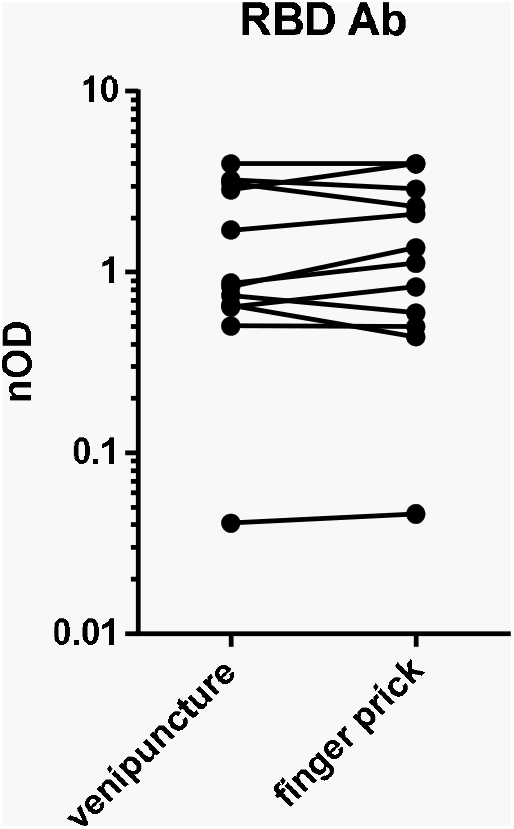
Comparison of venipuncture and finger prick. For the RBD-Ab we compared the antibody titers measured in eleven serum samples obtained from a venipuncture and that obtained by finger prick.

## Discussion

In this study, we describe the development of sensitive serological bridging assays based on the RBD and NP to detect antibodies against SARS-CoV-2. Assay performance was evaluated in serum samples of different patient populations (hospitalized and non-hospitalized) and the RBD bridging assay was found to best discriminate between cases and pre-outbreak controls across different time points, especially in individuals with COVID-19 symptoms who did not require hospital admission. We also show the early antibody kinetics in a group of non-hospitalized individuals, highly suspected of having contacted COVID-19, during their symptomatic phase. Finally, we demonstrate the feasibility of using finger prick sampling for the detection of antibodies against SARS-CoV-2.

In recent studies, seroconversion of SARS-CoV-2 infected hospitalized patients usually occurred between 5-14 days.[15,20,21,25,26] Our study demonstrated a similar result in non-hospitalized patients. Since an adequate antibody response requires time to develop, false-negative results will occur depending on the timing of sampling independent of the type of assay used, thereby influencing seroprevalence study results. In one patient followed over time, seroconversion occurred after day 15. Although this patient went to work (not healthcare related) and did groceries, other public measures regarding SARS-CoV-2 were followed, and this is probably a late seroconversion.

In the non-hospitalized patients, the isotype-specific antibody response to RBD follows a pattern similar to hospitalized patients [41], with a relatively late appearance of IgG antibodies. Although the IgG levels are significantly lower in the non-hospitalized population, these antibodies nevertheless are present in readily detectable amounts up to 60 days after symptom onset. The long-term persistence of these responses remains to be investigated.

Bridging assay, or homologous double-antigen sandwich immunoassays have found firm ground in the assessment of (weak) immune responses against biopharmaceutical drugs [27,28], but are as yet relatively uncommon for measurement of antibodies to viral antigens. Nevertheless, a commercially available assay using a similar format for SARS-CoV-2 has been described.[29] Advantages of this assay type include the ability to detect antibodies of any isotype (with the exception of IgG4 [30], which is unlikely to be of relevance in the context of an antiviral immune response) and a generally low background. Disadvantages include the lower responsiveness to low-affinity antibodies. In general, the assay described here is robust in detecting SARS-CoV-2 antibodies in patients with symptoms not requiring hospital admittance. This will aid in determining the true prevalence of COVID-19. The use of this developed assay could, therefore, lead to a better understanding of the dynamics of SARS-CoV-2 infections in the population since it is assumed that a large proportion of COVID-19 patients have mild symptoms and are not hospitalized.

In prior publications, mainly in hospitalized patients, the antibody response against the NP appeared earlier than that against the S protein. [18–23] Similarly, a study in SARS-CoV-1 described that the IgG immune response is more frequently directed against the NP compared to the S protein.[31] Moreover, in a SARS-CoV-2 study, it was found that most ICU patients had a higher N-IgG than S –IgG titer compared to non-ICU hospitalized patients.[32] Other studies demonstrated or suggested an association between disease severity and level and/or longevity of the antibody response for SARS-CoV-2 and MERS-CoV.[15,33–35] Our results suggest that the antibody response against the NP of SARS-CoV-2 needs more time to develop compared to the antibody response against the RBD in patients with mild symptoms, and is overall more variable. Our finding regarding the NP may, therefore, be (partially) based on the difference in immune response based on disease severity (e.g. hospitalized and non-hospitalized). However, the discrepancy between our finding and that of previous research needs to be confirmed in a larger sample size, since other factors such as the timing of samples and difference between assay types can also influence this.

Since different samples were positive in the RBD-Ab and NP-Ab assays in our healthy donors, there is a rationale for performing both assays to eliminate false positives. This will however result in a somewhat diminished sensitivity. A similar pattern is seen for the RBD-Ab assay compared to the RBD-IgG assay. In another study, combining different antigens has also been proposed to optimize the sensitivity and specificity of a SARS-CoV-2 serological assay.[23] Two-tier testing strategies are regularly implemented for other infectious diseases such as Lyme disease or syphilis.

In a recent study describing a SARS-CoV-2 transmission model a range of transmission scenarios were examined and it was suggested that: “serological testing is required to understand the extent and duration of immunity to SARS-CoV-2 which will help determine the post-pandemic dynamics of the virus.”.[36] Determining the true extent of the total incidence of SARS-CoV-2 by serological testing will optimize these transmission models and could aid in determining which public health measures are needed and their timing to maintain control of the SARS-CoV-2 pandemic. To assess this during periods of widespread transmission and when non-essential travel is prohibited, we developed a reliable serological assay which needs a low volume of serum and is therefore well suited to use for blood obtained by finger prick.

There are some limitations to this study. Further investigation of cross-reactivity for the different coronaviruses may be necessary since the RBD of SARS-CoV-2 may show cross-reactivity with other human coronaviruses (HCoV), and in particular SARS-CoV-1. However, we anticipate that this will not alter our findings due to the limited number of cases with SARS-CoV-1 in Europe. [37] By including a large population of healthy controls the risk of having missed substantial cross-reactivity against other HCoV is also limited; especially since antibodies against HCoV are detected in the majority of the population (>90% for all four HCoV).[38,39]

Furthermore, although this study provides an insight into the antibody response in the early phase of infection in non-hospitalized patients, the number of cases is still modest. Our study describes the early antibody kinetics in a population based on probable exposure and not based on symptoms or hospitalized patients who are RT-PCR positive. Yet, this provides an unbiased view of the performance of the developed serological assay and the antibody response in patients with mild SARS-CoV-2 infection.

In summary, this study demonstrates the use of a sensitive bridging assay to study seroprevalence in non-hospitalized COVID-19 patients, and provides insight into the early kinetics of the antibody response to SARS-CoV-2 in this population.

## Data Availability

Data from this study are available upon reasonable request

## Author contributions

GW, TR conceived of the study, EHV, FCL, BS, BJB, GW, TR were involved in study design and organization, NILD, SK, POH, GvM, FL, JYM, WL conducted experiments, EHV, FCL, TR analyzed data, EHV, FCL, GV, GW, TR wrote the paper, TR supervised the study, all authors provided critical revision of the paper.

---

**Figure S1.**
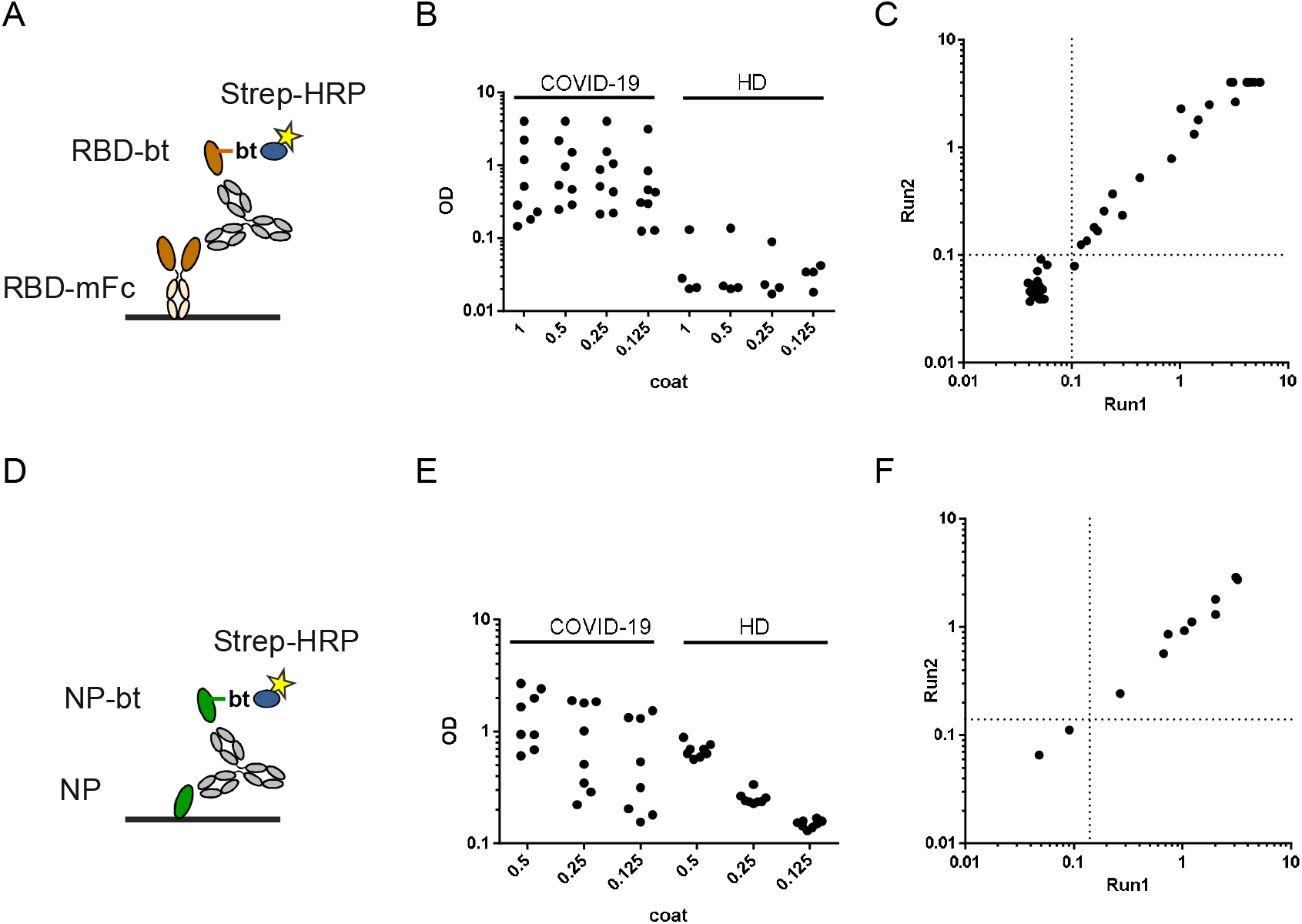
Setup of total antibody bridging assays for A) RBD and D) NP antigen. B,E) Impact of coating density on signals obtained from sera of selected COVID-19 patients and pre-outbreak controls for B) RBD, and E) NP. C,F) Correlation between two independent runs for C) the RBD-Ab assay, and F) the NP-Ab assay.

**Figure S2.**
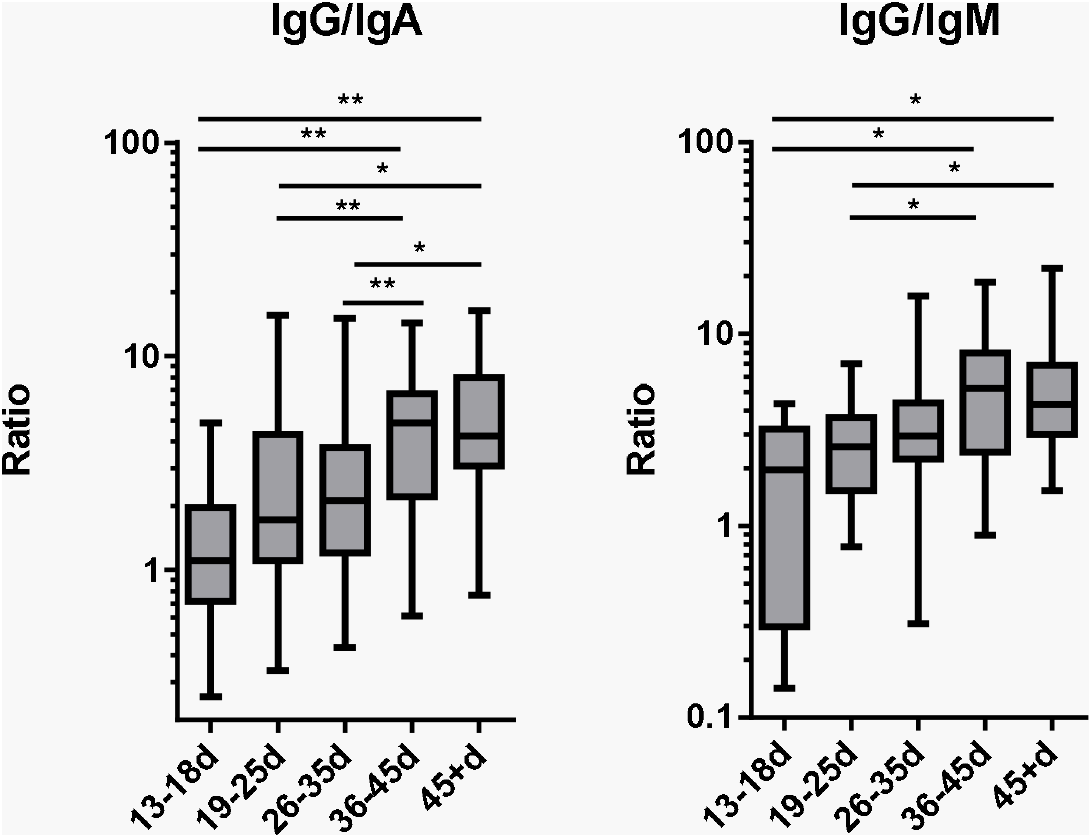
Gain in anti-RBD IgG over other isotypes over time in the non-hospitalized convalescent individuals. Ratio of nODs. * p < 0.05; ** p < 0.01; Kruskal-Wallis test/Dunn’s multiple comparisons test.

